# Accuracy of Tuberculosis Infection Diagnosis through IP-10-Based Assays for Immune Detection of Present Mycobacterium tuberculosis: A Cross-Sectional Evaluation

**DOI:** 10.64898/2026.02.24.26346999

**Authors:** Alexandru Stoichita, Madalina Burecu, Camelia Nita, Cristina Teleaga, Alexandru Daniel Radu, Mihaela Mihai, Beatrice Mahler, Elmira Ibraim

## Abstract

**Background:** Reliable detection of latent Mycobacterium tuberculosis (Mtb) infection (LTBI) remains challenging, particularly in TB contacts and immunocompromised individuals, where interferon-γ release assays (IGRAs) demonstrate variable sensitivity. IP-10, a chemokine produced at substantially higher concentrations than IFN-γ, represents a promising immune marker.

This study aimed to evaluate the diagnostic performance of two IP-10 based assays RIDA®QUICK TB (lateral flow) and RIDASCREEN® TB (ELISA), by comparison with QuantiFERON-TB Gold Plus (QFT-Plus) assay or a composite reference standard.

**Methods:** A cross-sectional diagnostic accuracy study enrolled 99 adults: 49 with culture-confirmed active pulmonary TB, 30 close TB contacts and 20 individuals with autoimmune disease, in Bucharest, Romania. All participants underwent RIDA Quick, RIDA Screen and QFT-Plus testing. Indeterminate results for all assays were reclassified using a composite reference standard.

**Results:** Against culture in active TB cases, RIDA®QUICK TB demonstrated a sensitivity of 85.7% (95% CI: 72.8–94.1) and PPV of 97.7%, while RIDA®SCREEN TB achieved 91.8% sensitivity (95% CI: 80.4–97.7) and 97.8% PPV. Specificity and NPV could not be reliably estimated due to the near-absence of true-negative individuals. Agreement with QFT-Plus was moderate to good (κ=0.47–0.93).ROC analysis performed against QFT-Plus as a comparator demonstrated good immunological discrimination for RIDA®QUICK TB (AUC = 0.828) and RIDA®SCREEN TB (AUC = 0.767), reflecting concordance with QFT-Plus rather than diagnostic accuracy against confirmed infection.

**Conclusion:** IP-10 based assays demonstrated higher sensitivity than QFT-Plus and excellent PPV across bacteriological standard, supporting their use as complementary tools for LTBI detection. Larger, more heterogeneous cohorts are needed to accurately define specificity and operational integration.

## Background

Respiratory pathogens are a significant contributor to illness and death, both in the European Union and worldwide, and will persistently pose a risk to public health during future outbreaks and global health crises (1). The COVID-19 pandemic underscored the pivotal role of diagnostics in monitoring and testing individuals irrespective of symptomatology, enabling timely public health interventions to curb viral transmission. Early identification of infection is essential to reduce both morbidity and mortality. Given the overlapping clinical presentation of respiratory infections, the use of rapid diagnostic methods is critical for accurate pathogen identification, guiding appropriate treatment strategies and containment measures. This is particularly relevant as respiratory infection syndromes account for a substantial proportion of deaths attributable to antimicrobial resistance worldwide (2).

Tuberculosis (TB), caused by Mycobacterium tuberculosis (Mtb), remains one of the world’s most lethal infectious diseases despite available preventive and curative therapies. In 2024, tuberculosis remained the leading cause of death from a single infectious agent worldwide. Globally, an estimated 10.7 million people developed TB, and approximately 1.23 million deaths were attributed to the disease, including around 150,000 deaths among people living with HIV, reflecting a modest decline compared with 2023 but a burden that remains unacceptably high (3). In the WHO European Region, sustained progress has been observed over the past decade. According to the ECDC/WHO Tuberculosis Surveillance and Monitoring in Europe 2025 report, EU/EEA countries reported 38,993 TB cases in 2023, corresponding to a notification rate of 8.6 cases per 100,000 population, while TB-related mortality remained low at the regional level. Compared with 2015, the WHO European Region has achieved an estimated 39% reduction in TB incidence and a 49% reduction in TB deaths, although substantial heterogeneity persists between countries (3,4).

Romania continues to report the highest tuberculosis burden among European Union (EU) member states, despite a sustained decline in TB incidence and mortality over the past decade. According to the WHO Global Tuberculosis Report 2025, Romania is classified as a lower-moderate TB incidence country, yet its TB notification rates remain substantially higher than the EU/EEA average, which is low and relatively stable. This pattern is corroborated by the ECDC/WHO Tuberculosis Surveillance and Monitoring in Europe 2025 report, which documents an EU/EEA notification rate of 8.6 cases per 100,000 population in 2023, while Romania continues to contribute a disproportionate share of notified TB cases, including the highest notification rates among young children, indicative of ongoing transmission. Clinically, TB-related mortality in Romania remains several-fold higher than the EU average, suggesting delayed diagnosis, advanced disease at presentation, and the persistent influence of social vulnerability and comorbidities. Although Romania follows the overall downward epidemiological trend observed in the WHO European Region, these indicators highlight persistent clinical and epidemiological disparities within the EU, underscoring the need for strengthened early case detection, targeted interventions in high-risk populations, and improved continuity of care to further reduce TB burden and mortality (3,4).

The WHO End TB Strategy aims to eliminate TB by reducing incidence, mortality, and associated suffering, with a major pillar focused on expanding access to preventive treatment for high-risk individuals. Latent TB infection (LTBI) represents a sustained immune response to Mtb antigens in the absence of clinical disease. It is estimated that 25% of the global population is infected. Accurate detection of LTBI is essential to identify individuals who may benefit from TB preventive therapy and thereby interrupt progression to active disease (5).

Currently, there are two classes of tests available for LTBI: interferon-gamma release assays (IGRAs) and the tuberculin skin test (TST). These tests are indirect and rely on a strong immune response to accurately identify individuals who are infected with TB(6).

The TST, although inexpensive and widely available, suffers from limited specificity due to BCG vaccination and environmental non-tuberculous mycobacteria (NTM), and from reduced sensitivity in immunocompromised individuals (6).

IGRAs, including QuantiFERON-TB Gold Plus (QFT-Plus) and T-SPOT.TB, offer greater specificity by measuring IFN-γ responses to Mtb–specific antigens and are unaffected by BCG or most NTM. However, IGRAs remain more costly, require stricter pre-analytical handling, cannot distinguish infection from active disease, and may yield false-negative or indeterminate results in early TB, low-burden infection, or immune-dysregulated hosts (7)(8).

IP-10 is a highly promising complementary biomarker for LTBI diagnosis. Although induced downstream of IFN-γ, it is produced in substantially higher concentrations by antigen-presenting cells and can also be stimulated by IL-2, type I IFs, IL-27, IL-17, IL-23, TNF-α and IL-1β. Its markedly higher dynamic range and stable signal-to-background ratio allow more robust detection using ELISA or lateral-flow platforms compared with IFN-γ, supporting IP-10 as a reliable indicator of antigen-specific cell-mediated immune activation (9–15).

The utilization of assays that evaluate IP-10 could have the potential to enhance LTBI detection. In the current study we aimed to use such assays using the RIDA®QUICK TB and the RIDASCREEN® TB tests.

Currently, there are few published studies that assess the diagnostic accuracy of RBAG developed tests, specifically in contrast to the QFT-Plus assay. Diagnostic accuracy studies are crucial for evaluating new diagnostic methods against the target product profiles set by the World Health Organization and for comparing them to existing diagnostic procedures.

**Aim** of this study was to narrow the gap in knowledge by collection of data on the diagnostic accuracy of the RBAG-developed tests in 3 group of individuals: with active TB disease, TB contacts and those with an autoimmune disease. The accuracy of the RBAG test was compared with that of the QFT-Plus assay.

## Objectives

### Primary objective

To evaluate the accuracy of two tests, one ELISA and one lateral-flow, based on IP-10 detection, for the diagnosis of LTBI.

### Secondary objectives

1. To assess the LTBI diagnostic performance of the RBAG developed tests in persons with presumptive LTBI (TB contacts), through comparison with QFT-Plus assay
2. To identify the individual demographic and clinical factors impacting the results of the tests in subjects with active TB disease confirmed by positive Mtb culture
3. To analyze the LTBI diagnostic capacity of RBAG tests versus QFTB-Plus assay, in subjects with an autoimmune disease

### Study design and setting

This study is a cross-sectional diagnostic performance evaluation study in 3 groups of subjects to mitigate bias in the estimation of clinical performance characteristics, conducted in a TB dispensary of The Marius Nasta Institute of Pneumophtiziology in Bucharest, Romania.

## Material and methods

### Study population

The study intended to include 100 patients, aged 18 years or older, divided into 3 groups: one group of 50 patients with active pulmonary TB disease, one other of 30 patients with presumptive LTBI (TB contacts) and the third of 20 patients with an autoimmune illness.

Given the absence of robust prior data for IP-10 assays performance, a formal power calculation was not feasible; a target sample of 100 participants was selected for exploratory evaluation of diagnostic accuracy.

### Inclusion criteria

- adults (aged 18 years or older)
- with active pulmonary TB confirmed by positive Mtb culture from sputum (group 1)
- close contact of an individual with active pulmonary TB and positive microscopy of sputum (group 2)
- with a documented autoimmune disease diagnostic (group 3)
- provided consent to participate in the study, after being fully informed.

### Exclusion criteria

- minor (below 18 years old)
- with active pulmonary TB, but negative culture for Mtb from sputum (group 1)
- no documented close contact of an individual with active TB positive for Mtb in sputum microscopy (group 2)
- no documented diagnostic of an autoimmune disease (group 3)
- treatment for TB disease or LTBI completed within the past 12 months
- any medication with activity against Mtb taken within two weeks before study entry
- not able or not willing to consent to participate in the study

### Principles of the tests

Prior to use of the RIDA®QUICK TB test, the (whole) blood samples must be pre treated using the RIDA® TB Tubes to induce the analyte IP-10. This is done either by direct blood collection into each of the three RIDA® TB Tubes or alternatively by blood collection into a lithium heparin tube (indirect blood collection) with subsequent transfer of the blood into each of the three RIDA® TB Tubes. This is followed by incubation and plasma collection. The pre-treatment is performed in accordance with the instructions for use of the RIDA® TB Tubes. For each tested person, three plasma samples must be analyzed, to obtain a valid test result (RIDA® TB Tubes NEG, TEST, POS). The RIDA®QUICK TB test is a one-step immunochromatographic lateral flow test, detecting IP-10 by the formation of an antibody-antigen sandwich. If IP-10 is present in one of the three plasma samples that have to be tested per person (RIDA® TB Tubes NEG, TEST, POS), an antibody-antigen sandwich is formed and bound to the test line. The antibody-antigen sandwich is visualized by the use of labelled colloidal gold nanoparticles and is manifested by a red violet coloration at the test line. Non complexed gold-labelled antibodies are bound to the subsequent control line, which is manifested by a red violet coloration at the control line. In negative samples, gold labelled immune complexes do not bind to the test line; only the control line is formed. The red violet control line always indicates if the test is valid. The generated signal is measured using the RIDA®Q3 and converted into the corresponding IP-10 sample concentration using a standard curve stored in the device.

Before using the RIDASCREEN® TB test, the (whole) blood samples must be pretreated using the RIDA® TB Tubes Tubes to induce the analyte IP-10. This is done either by direct blood collection into each of the three RIDA® TB Tubes or alternatively by blood collection into a lithium heparin tube (indirect blood collection) with subsequent transfer of the blood into each of the three RIDA® TB Tubes. This is followed by incubation and plasma collection. The pre-treatment is performed in accordance with the instructions for use of the RIDA® TB Tubes. The RIDASCREEN® TB test, employs specific antibodies in a sandwich method. Monoclonal antibodies against IP-10 are bound to the well surface of the microtiter plate. The plasma samples (three per person to be tested - RIDA® TB Tubes NEG, TEST, POS) as well as the standards and controls are pipetted into the wells of the microtiter plate together with biotinylated monoclonal anti-IP-10 antibodies (Conjugate 1) for incubation at room temperature (20 - 25 °C). After a washing step, streptavidin-poly-peroxidase conjugate (Conjugate 2) is added and incubated again at room temperature (20 - 25 °C). If IP-10 is present in the plasma samples, a sandwich complex forms, which consists of immobilized antibodies, IP-10 and the antibodies conjugated with the biotin-streptavidin peroxidase complex. A further washing step removes the unbound streptavidin-poly-peroxidase. After the addition of substrate to positive samples, the bound peroxidase converts the substrate into a chromogenic compound, causing a blue color reaction in the wells of the microtiter plate. The addition of a stop reagent changes the color from blue to yellow. The absorbance is proportional to the concentration of IP-10 present in the plasma sample.

### Reference standards

Mtb culture from sputum was defined as the primary microbiological reference standard. GeneXpert and sputum smear microscopy served as secondary bacteriological references. QFT-Plus was treated as a comparator immunological assay rather than a diagnostic gold standard.

### Handling of indeterminate results

Indeterminate results for RIDA®QUICK TB RIDASCREEN® TB and QFT-Plus were reclassified using a composite reference standard of Mtb from sputum: GeneXpert test, smear microscopy and culture. Indeterminate results in confirmed TB cases were coded as positive, reflecting expected immune activation.

Indeterminate results in TB contacts and subjects with autoimmune disease, without bacteriological or radiological evidence of active TB, were coded as negative.

This reclassification strategy was applied solely to minimize analytical exclusions and was not intended to redefine true infection status, acknowledging the potential for incorporation bias inherent to disease-enriched diagnostic studies.

## Data collection, management and analysis

### Data sources

Study data were systematically recorded in a secure electronic database. Source documentation included the following:

- Medical records for demographic and clinical variables
- Registers for TB case notification
- TB laboratory registers for results of microbiological and molecular assays from sputum
- Immunology laboratory registers for tests results

### Data analysis

The clinical and laboratory investigators reviewed, analyzed, cleaned the clinical data into the data source documents and uploaded them into the secure electronic database.

Then, the data have been examined using suitable statistical software, such as Stata or R. Sensitivity, specificity, positive predictive value (PPV), negative predictive value (NPV), accuracy, and κ coefficient were calculated with 95% confidence intervals. ROC curve analysis was predefined to evaluate immunological discrimination and concordance between IP-10–based assays (RIDA®QUICK TB and RIDASCREEN® TB) and the QFT-Plus comparator, rather than diagnostic accuracy against microbiologically confirmed infection. This approach was selected a priori because the study cohort, particularly the active TB subgroup, was disease-enriched and lacked a sufficient number of true-negative reference cases required for valid specificity-driven ROC analyses against bacteriological standards. Consequently, ROC-derived AUC values reflect agreement with QFT-Plus–defined immune status rather than confirmation of true infection.

## Results

### Study population and demographic characteristics

A total of 100 adults were enrolled and allocated into 3 predefined groups: subjects with active pulmonary TB (n=50), close contacts of individuals with microscopy-positive pulmonary TB (n=30) and subjects with an autoimmune disease undergoing evaluation for LTBI (n=20).

One participant in group 1, even positive for GeneXpert test and microscopy from sputum, did not find the inclusion criteria, being that the culture examination did not show growth. Therefore, he was eliminated from the statistical analysis, which has been done on just 49 cases in group 1 and on 99 study subjects in total.

All other participants met the eligibility criteria and none of exclusion criteria. No protocol deviation was recorded.

Participant’s age ranged from 23 to 90 years. Median age increased progressively across groups, from 45.04 years in the active TB group to 46.50 years among TB close contacts and 53.75 years within the cohort with autoimmune disease. The active TB group demonstrated the widest distribution, including a single 90-year-old outlier. (Tab. 1)

**Table 1.**
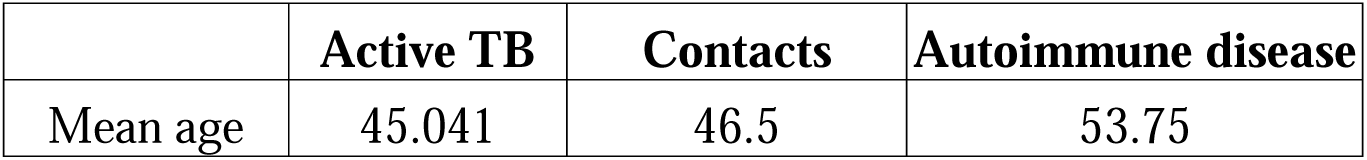
Distribution by age.

|  | **Active TB** | **Contacts** | **Autoimmune disease** |
| --- | --- | --- | --- |
| Mean age | 45.041 | 46.5 | 53.75 |

Overall, 55% of the total was female. Gender distribution varied markedly between groups: the active TB group was predominantly male (73%), all close contacts were female, and the autoimmune cohort showed a near-balanced sex ratio (55% female). (Fig. 1)

**Fig. 1.**
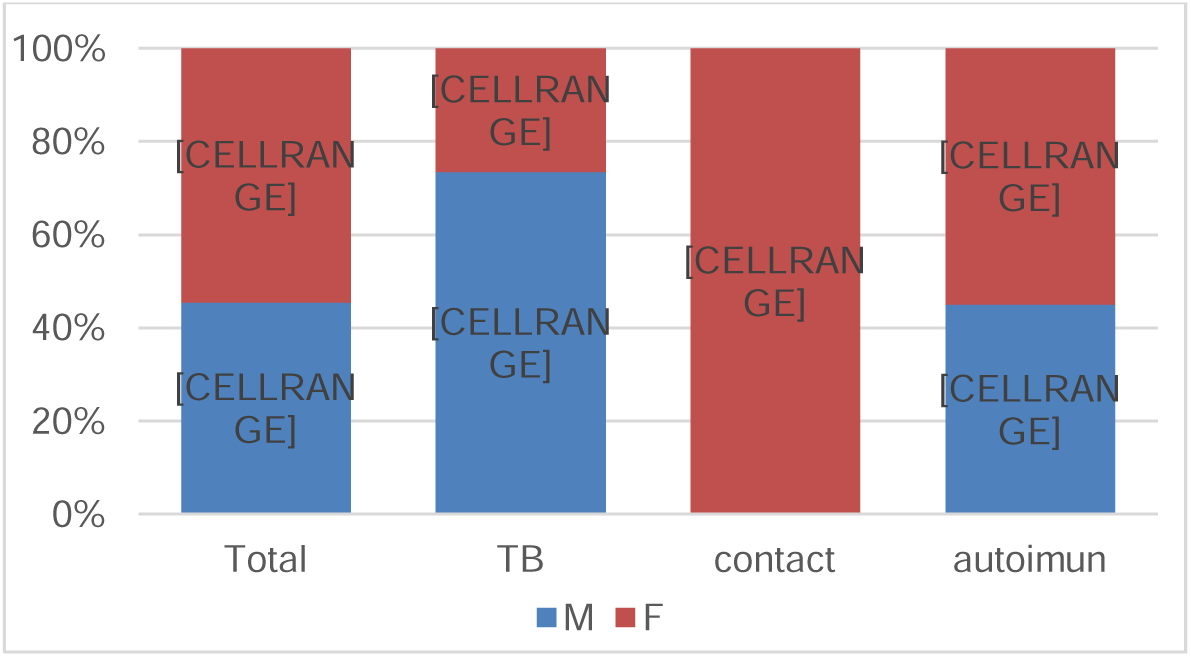
Structure of groups by gender

### Residential and behavioral characteristics

Residential status was predominantly urban, with 82% of participants residing in urban area. The distribution was similar across study groups, ranging from 78% (in active TB group) to 90% (in TB contacts), indicating no major imbalance in recruitment by residential setting. (Fig. 2)

**Fig. 2.**
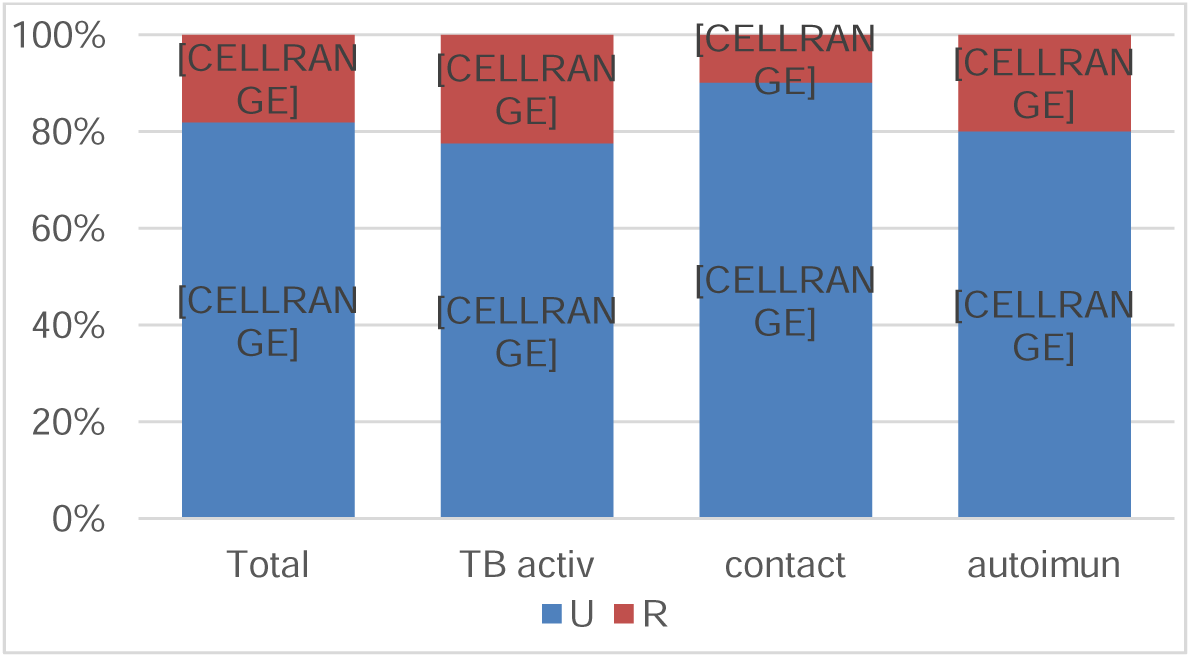
Structure of groups by residence

Regarding smoking status, 52% of participants were non-smokers, 40% were current smokers and 8% were former smokers. The highest proportion of current smokers was observed in individuals with active TB – 55%. (Fig. 3)

**Fig. 3.**
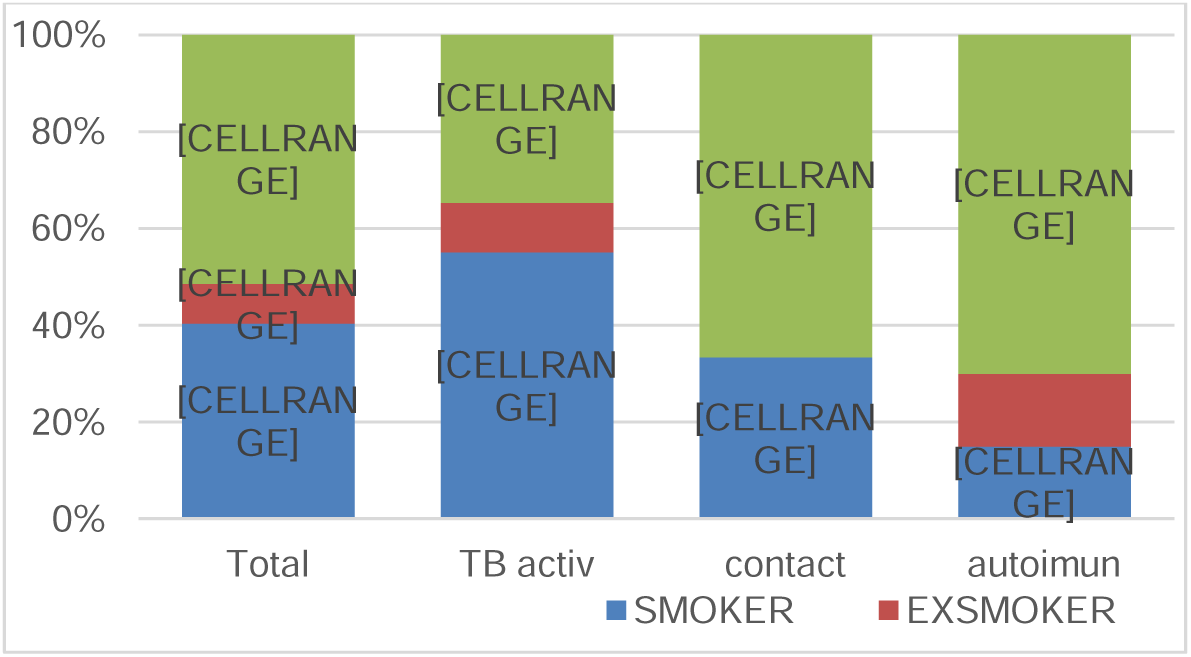
Distribution of groups by smoking status

### Diagnostic performance of IP-10 based assays compared with QFT-Plus

In our cohort both RIDA®SCREEN TB and RIDA®QUICK TB assays demonstrated a superior ability to detect TB infection among individuals with active TB disease. The performance of the two IP-10 based assays was largely comparable, with both showing a consistent trend toward a higher proportion of positive results relative to the QFT-Plus assay (Fig 4). This pattern was confirmed across concordance and sensitivity analyses, suggesting that IP-10 mediated immune responses may offer improved diagnostic responsiveness in active TB.

**Fig. 4.**
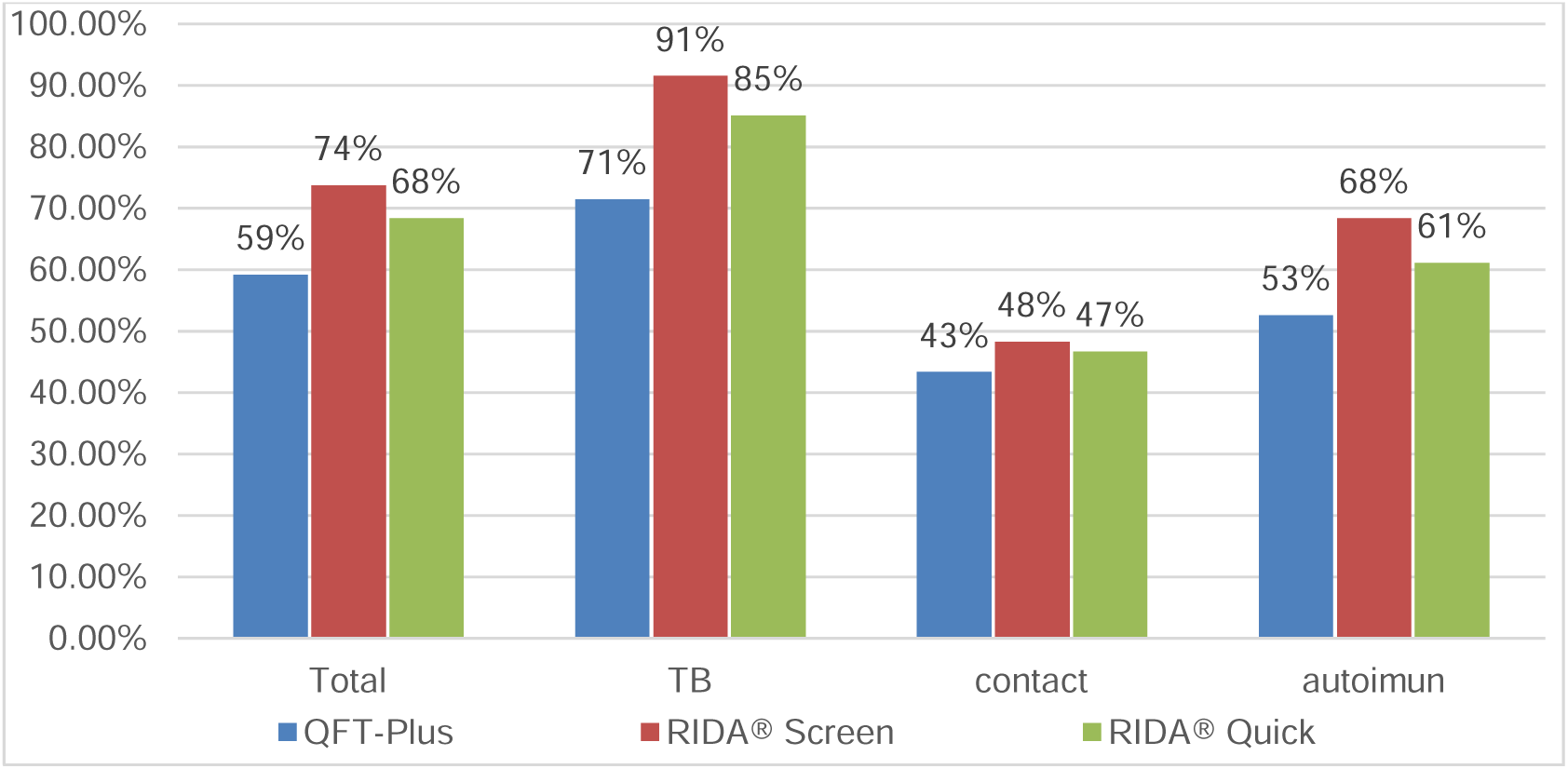
Positivity rate by study group and test type

#### A. Overall test positivity and microbiological reference comparison

A total of 100 participants were enrolled in the study. Complete paired results for RIDA®QUICK TB, RIDASCREEN® TB, and QuantiFERON-TB Gold Plus (QFT-Plus) were available for 99 individuals and were included in the comparative analyses.

In the subgroup of participants with culture-confirmed active pulmonary TB (N = 49), the proportion of culture-positive cases detected by each assay is summarized in Table 1. Both IP-10–based assays identified a higher number of culture-confirmed cases compared with QFT-Plus, with sensitivities of 85.7% for RIDA®QUICK TB and 91.8% for RIDASCREEN® TB, versus 71.4% for QFT-Plus. As this subgroup contained virtually no culture-negative individuals, specificity and negative predictive value could not be reliably estimated.

Overall test positivity across all study groups is presented in Table 2. The table summarizes the number of positive results obtained with each assay among participants with active TB, TB contacts, and individuals with autoimmune disease, as well as in the total cohort with complete paired results (N = 99). Across all groups, both IP-10–based assays yielded a higher number of positive results than QFT-Plus, with the largest differences observed in the active TB subgroup.

**Table 1:**
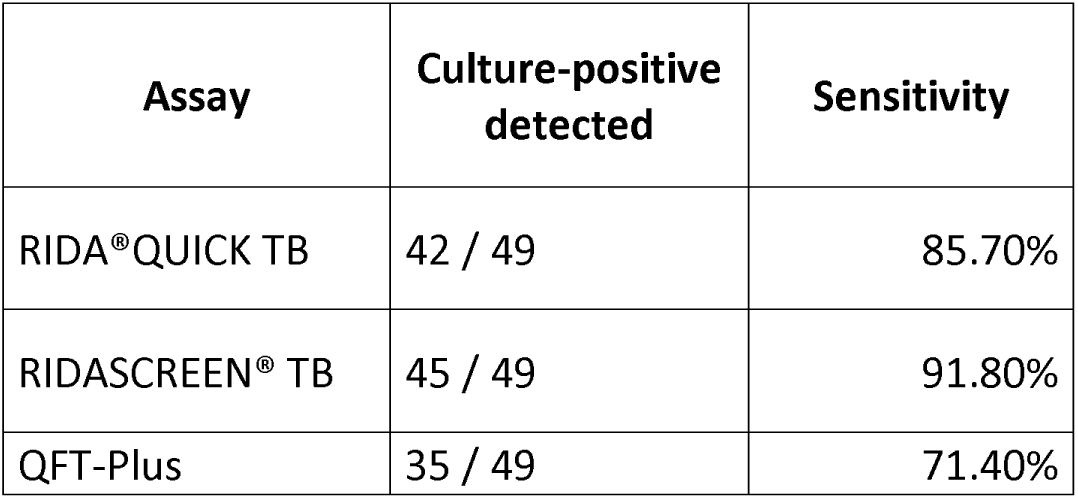
Diagnostic performance in culture-confirmed active TB.

**Table 2:** Test positivity in TB contacts and autoimmune diseases.

| **Study group** | **N** | **RIDA®QUICK TB positive** | **RIDASCREEN® TB positive** | **QFT-Plus positive** |
| --- | --- | --- | --- | --- |
| TB contacts | 30 | 14 / 30 | 14 / 30 | 13 / 30 |
| Autoimmune disease | 20 | 11/20 | 13 / 20 | 10/20 |
| **Total (complete paired results)** | **99** | **67 / 99** | **72 / 99** | **58 / 99** |

#### B. Comparative diagnostic accuracy of RIDA®QUICK TB and QFT-Plus (Annex 1)

In the full cohort, RIDA®QUICK TB showed good overall agreement with QFT-Plus (κ=0.677), although the McNemar test (p=0.035) demonstrated a significant directional disagreement, with RIDA®QUICK TB classifying more samples as positive (12 vs. 3).

In discordant pairs, RIDA®QUICK TB produced a positive result more frequently than QFT-Plus, suggesting detection of immune activation not captured by IFN-γ–based readouts. The assay showed highPPA (∼95%), NPV=0.906, and PPV=0.821, while NPV was moderate (70.7%), consistent with an expected shift toward increased positivity in an IP-10–driven assay.

##### a. Subgroup of subjects with active TB disease

In individuals with active TB, agreement between RIDA®QUICK TB and QFT-Plus was moderate (κ=0.471), with an overall accuracy of 81.6%. PPAremained very high (97.1%) and PPV substantial (0.810). NPA, however, was low (0.429), driven by the higher number of RIDA®QUICK TB positive/QFT-Plus negative results (8 vs. 1). The McNemar test (p=0.039) confirmed a statistically significant directional imbalance. Importantly, because QFT-Plus is not a definitive reference standard for active TB, these apparent false positives represent true antigen-specific immune responses, particularly in early disease, low bacillary burden, or fluctuating IFN-γ production.

##### b. Subgroup of subjects with TB contact

Among close contacts of TB cases, agreement between the assays was almost perfect (κ=0.933), with an accuracy of 96.7%. PPA and NPV were 1.0 each, while NPA(0.941) and PPV (0.929) were similarly high. No directional disagreement was observed (McNemar p=1.00). These results reflect excellent assay concordance in a lower-risk population, although interpretation should consider the modest sample size.

##### c. Subgroup of subjects with autoimmune disease

In participants with autoimmune disease, agreement was moderate (κ=0.500), with an overall accuracy of 75%. PPA(0.800) and NPV (0.778) were acceptable, while NPA(0.700) and PPV (0.727) were lower than in the other subgroups. No directional asymmetry was identified (McNemar p=1.00). The more variable performance in this subgroup likely reflects underlying immune dysregulation, which can differentially affect IFN-γ release and IP-10 response.

## ROC curve analysis

ROC curve analysis was performed to evaluate the degree of immunological discrimination and concordance between IP-10–based assays and the QFT-Plus comparator, rather than to estimate diagnostic accuracy against confirmed infection. This approach was adopted because the study cohort, particularly the active TB subgroup, was disease-enriched and lacked an adequate number of true-negative individuals required for specificity-driven diagnostic ROC analyses against microbiological reference standards.

In the overall cohort, ROC analysis demonstrated good discriminatory performance of RIDA®QUICK TB relative to QFT-Plus, with an AUC of 0.828 (95% CI: 0.736–0.919). Subgroup analyses revealed marked variability across the three clinical categories. Among close TB contacts, discrimination was excellent (AUC = 0.971), consistent with the high level of concordance observed between the two assays in this group. In contrast, among individuals with active TB, discriminatory performance was moderate (AUC = 0.700).

In participants with autoimmune disease, discriminatory ability was acceptable (AUC = 0.750); however, the wide confidence intervals indicate limited precision, reflecting both the small sample size and the expected variability associated with immune dysregulation.

Importantly, these ROC curves reflect agreement with QFT-Plus rather than confirmation of true infection status. Because QFT-Plus is not a definitive diagnostic reference for active TB, the reported AUC values represent immunological concordance between assays rather than diagnostic accuracy against microbiologically confirmed disease. Accordingly, interpretation of ROC-derived metrics should be integrated with bacteriological reference standards and clinical context across the cohort.

### C. Comparison of RIDA®QUICK TB with microbiological reference standard in the group of active TB disease subjects (Annex 2)

Sputum culture was positive in 49 of the 50 patients with active pulmonary TB (98%). RIDA®QUICK TB identified 42 of these 49 culture-positive cases, corresponding to a sensitivity and an overall accuracy of 85.7% (42/49) each.

Because there were no culture-negative samples within the N=49 comparison set (i.e., 0 true negatives), specificity and NPV cannot be validly estimated (they are undefined/non-informative in this context). Consequently, any values for these parameters, as well as a near-zero kappa, primarily reflect extreme class imbalance and the absence of true-negative reference observations, rather than genuine disagreement between tests. The McNemar test indicated a statistically significant directional discordance (χ²=5.14; p=0.023), with more culture positive/RIDA®QUICK TB negative pairs than the reverse (7 vs. 0). This result is consistent with a modest asymmetry in discordant classifications, suggesting that RIDA®QUICK TB missed a small number of culture-positive cases in this highly disease-enriched subgroup. However, interpretation should remain cautious given the very small number of discordant pairs, the absence of culture-negative reference samples, and the resulting extreme class imbalance, which limits broader conclusions about systematic bias. (Tab. 2)

**Table 2.** Comparison of RIDA®QUICK TB with microbiological reference standard.

| **N=49** | **Sensitivity** | **Specificity** | **PPV** | **NPV** | **Accuracy** |
| --- | --- | --- | --- | --- | --- |
| RIDA®QUICK TB vs<br>Mtb culture | 0.857 | - | 1.000 | 0.000 | 0.857 |

### D. Comparative diagnostic accuracy of RIDA®SCREEN TB and QFT-Plus (Annex 3)

RIDA®SCREEN TB showed good agreement with QFT-Plus (κ=0.562), through McNemar test (p=0.0036), indicating a directional imbalance driven by more RIDA®SCREEN TB positive classifications (17 vs. 3). The assay demonstrated high sensitivity (∼95%), moderate specificity (58.5%) and strong predictive performance (PPV=0.764; NPV=0.889).

**a.** In **the active TB subgroup** (N=49), agreement was moderate (κ=0.365; accuracy 80%). RIDA®SCREEN TB showed sensitivity of 91.83% and NPV of 1.0, PPV=0.783 and low specificity (0.286), reflecting more positive classifications when QFT-Plus was negative. The McNemar test (p=0.002) indicated significant directional disagreement. Given that QFT-Plus is not a definitive standard test for active disease, some discordant results may represent true biological response rather than false positive.
**b.** In **TB contacts** (N=30), agreement was very good (κ=0.798; accuracy 90%), with high sensitivity (0.923), specificity (0.882), PPV (0.857) and NPV (0.938). No directional asymmetry was observed (McNemar p=1.00).
**c.** In **subjects with autoimmune disease** (N=20), agreement was poor-moderate (κ=0.300; accuracy 65%). Sensitivity was 0.800, specificity 0.500, and PPV/NPV was 0.615/0.714. No significant disagreement was detected (McNemar p=0.453). Some divergence from QFT-Plus is expected given underlying immune dysregulation.

Across subgroups interpretation should remain cautious, due to the limited samples size, which may influence the stability of agreement and performance estimates.

Compared with QFT-Plus, RIDA®SCREEN TB demonstrated high sensitivity and strong NPV, with a tendency to yield more positive classifications in discordant pairs, most evident in the active TB subgroup. Agreement was highest in TB contacts, moderate in active TB cases, and more variable in those with autoimmune disease, reflecting expected immunological heterogeneity. As with RIDA®QUICK TB, PPV and NPV reflect the underlying distribution of QFT-Plus positivity in the sample and should not be interpreted as performance against confirmed disease. For clinical interpretation, correlation with bacteriological reference standard (Mtb culture) remains essential.

In the overall cohort (N=99), RIDA®SCREEN TB showed good discriminatory ability relative to QFT-Plus (AUC=0.767; 95% CI: 0.664-0.869; p<0.001), indicating reasonable separation between QFT-Plus positive and QFT-Plus negative cases. Discrimination was excellent in contacts (AUC=0.903), while performance across other subgroups was heterogeneous, consistent with the RIDA®QUICK TB/QFT-Plus comparison.

### D. Comparison of RIDA®SCREEN TB with microbiological reference standard in the group of active TB disease subjects (Annex 4)

Mtb culture from sputum was positive in 49 of 50 individuals (98%). RIDA®SCREEN TB correctly identified 45 of these culture-positive cases, yielding a sensitivity and an overall accuracy of 91.8% each. Because the subgroup contained no culture-negative reference samples (0 true negatives), specificity cannot be estimated, and NPV is not informative in this setting. Consequently, agreement statistics (i.e., κ∼0) and low/zero values for specificity/NPV primarily reflect extreme class imbalance and the absence of reference negatives, rather than a true limitation of assay performance.

Importantly, the McNemar test showed no significant directional discordance vs culture (χ²=2.250; p=0.134; discordant pairs 4 vs 0), indicating no evidence of systematic bias in discordant classifications. (Tab. 3)

**Table 3.** Comparison of RIDA®SCREEN TB with microbiological reference standard.

| **N=49** | **Sensitivity** | **Specificity** | **PPV** | **NPV** | **Accuracy** |
| --- | --- | --- | --- | --- | --- |
| RIDA®SCREEN TB vs<br>Mtb culture | 0.918 | - | 1.000 | 0.000 | 0.918 |

### E. Comparison of QFT-Plus with microbiological reference standard in the group of active TB disease subjects (Annex 5)

QFT-Plus identified 35 of the 49 culture-positive TB cases, corresponding to a sensitivity of 71.4%. In this subgroup, all culture results were positive (no reference negatives); therefore, specificity and NPV cannot be meaningfully estimated, and the PPV is effectively 100% (35/35), because there were no culture-negative samples to generate false-positive QFT-Plus results. The McNemar test showed significant directional discordance (χ²=12.07; p=0.00051), driven by a clear imbalance in discordant pairs (14 culture positive/QFT-Plus negative vs 0 culture negative/QFT-Plus positive), indicating that QFT-Plus failed to detect a substantial proportion of culture-confirmed cases in this cohort.

## Discussions

Across the study cohort, both IP-10–based assays demonstrated higher sensitivity than QFT-Plus, with RIDA®QUICK TB consistently classifying a greater number of participants as positive, particularly among individuals with active TB or immune dysregulation. This pattern reflects the biological nature of the assays: IP-10 and IFN-γ release measurements capture antigen-specific immune activation rather than direct detection of viable organisms. Importantly, higher positivity rates observed with IP-10–based assays compared with QFT-Plus should not be interpreted as false positivity but rather as detection of antigen-specific immune activation not consistently captured by IFN-γ–only platforms. Consequently, predictive values depend on the underlying prevalence of immunological sensitization and should not be interpreted as direct proxies for true infection. For this reason, comparison with microbiological standard (Mtb culture) is essential to determine whether additional RIDA® positive results represent early immunological responses, low-burden infection, or assay discordance.

In the bacteriologically confirmed active TB subgroup (N=49; all sputum cultures positive), RIDA®QUICK TB identified 42 of 49 culture-positive cases, corresponding to a sensitivity of 85.7%, a PPV of 100% (42/42), and an overall accuracy of 85.7%. Specificity, NPV, and κ values could not be reliably estimated due to the near absence of culture-negative individuals in this disease-enriched cohort, resulting in numerically low and statistically unstable estimates driven by extreme class imbalance rather than true analytical disagreement.

RIDA®SCREEN TB showed comparable diagnostic performance. In the active TB group, sensitivity remained high (91.8%), PPV 100% and accuracy 91.8%, against bacteriological reference, as RIDA®SCREEN TB specificity and NPV could not be interpreted due to the absence of true-negative samples. RIDA®SCREEN TB identified 45 of 49 culture-positive cases, with no significant directional discordance. Instances in which RIDA®SCREEN TB or RIDA®QUICK TB were positive while culture results were negative are biologically plausible: IP-10 reflects cellular immune sensitization, whereas cultures depend on bacillary load and viability. Such discrepancies may arise in early infection, low-burden disease, heterogeneous sputum sampling, or temporal differences between immune activation and bacteriological detectability.

The findings of this study are consistent with emerging external evidence supporting the diagnostic value of IP-10–based assays. A multicenter evaluation by Köffer et al. (16) reported higher sensitivity for both RIDA®SCREEN TB and RIDA®QUICK TB compared with QFT-Plus, along with strong PPV performance against a composite clinical reference closely matching the performance patterns observed in our cohort. Likewise, recent reviews, including the analysis by Alonzi et al. (17), have identified RIDA® QUICK TB as part of a new generation of IP-10–based IGRAs with improved sensitivity relative to IFN-γ–only platforms. The superior sensitivity of both IP-10 assays observed in our study aligns with these findings and supports the growing evidence that IP-10 may provide a more robust immunological signal in active and incipient TB. Taken together, the results of these studies highlight that although the dataset’s disease-enriched composition enables precise estimation of sensitivity and PPV, it does not allow reliable calculation of specificity or NPV, which require adequate representation of true-negative individuals. A more complete assessment of diagnostic performance particularly for specificity will require validation in more balanced populations such as close contacts, immunocompromised individuals, and low-risk clinical groups, as well as the application of composite clinical reference standards across the full cohort. These findings underscore the potential of IP-10 assays as valuable complementary tools within TB diagnostic pathways and support further investigation into their utility in diverse epidemiological and immunological contexts.

## Limitations

This study has several limitations. The subgroup with active TB was disease-enriched, preventing reliable estimation of specificity and NPV. The subgroup with autoimmune diseases was small and various degrees of immune suppression may have influenced test performance. All participants were adults and most of them from urban area of Bucharest, limiting generalizability to rural, pediatric or HIV-infected populations. ROC analyses were performed against an immunological comparator rather than a definitive diagnostic gold standard, limiting inference regarding specificity-driven diagnostic accuracy.

Finally, longitudinal follow-up was not performed, preventing evaluation of PPV for progression.

## Clinical implications

The high sensitivity and consistently strong PPVs observed for both IP-10 assays underscore their potential utility as complementary diagnostic tools, particularly in settings where rapid immune-based testing is needed to support early clinical decision-making. Their ability to detect immunological responses, even when conventional assays underperform, suggests a valuable role in identifying low-burden or early disease.

Future multicenter studies involving more heterogeneous and representative populations, together with longitudinal follow-up, are essential to accurately define specificity, evaluate predictive performance over time and determine how IP-10–based assays may be integrated into broader LTBI diagnostic pathways and preventive therapy algorithms.

## Conclusion

In this diagnostic tools evaluation, the IP-10–based RIDA®QUICK TB and RIDA®SCREEN TB assays demonstrated high sensitivity and strong PPV performance, outperforming QFT-Plus in detecting immune response associated with Mtb infection. RIDA®SCREEN TB achieved the highest sensitivity against culture, supporting the premise that IP-10–mediated response may better capture early or low-burden disease where IFN-γ–based assays often show reduced yield.

The low estimates of specificity and NPV observed in the active TB subgroup reflect the absence of true-negative individuals in this disease-enriched cohort, rather than limitations of the assays themselves. When interpreted alongside clinical, imagistic and microbiological data, IP-10 assays may help reduce missed infections and facilitate earlier treatment decision.

These findings align with growing multicentre evidence identifying IP-10 as a promising next-generation biomarker. Further studies in more diverse and representative populations are needed to fully characterize specificity, NPV and operational applicability. Nonetheless, the present results support the integration of RIDA®QUICK TB and RIDA®SCREEN TB as complementary tools within TB infection diagnostic pathways, with potential advantages for early detection and improved patient management.

## Data Availability

All data produced in the present study are available upon reasonable request to the authors

## ANNEX 1 Statistical analysis

**1. Comparative Diagnostic Accuracy of RIDA Quick and QFT Stratified by Clinical Category (ANNEX 1)**

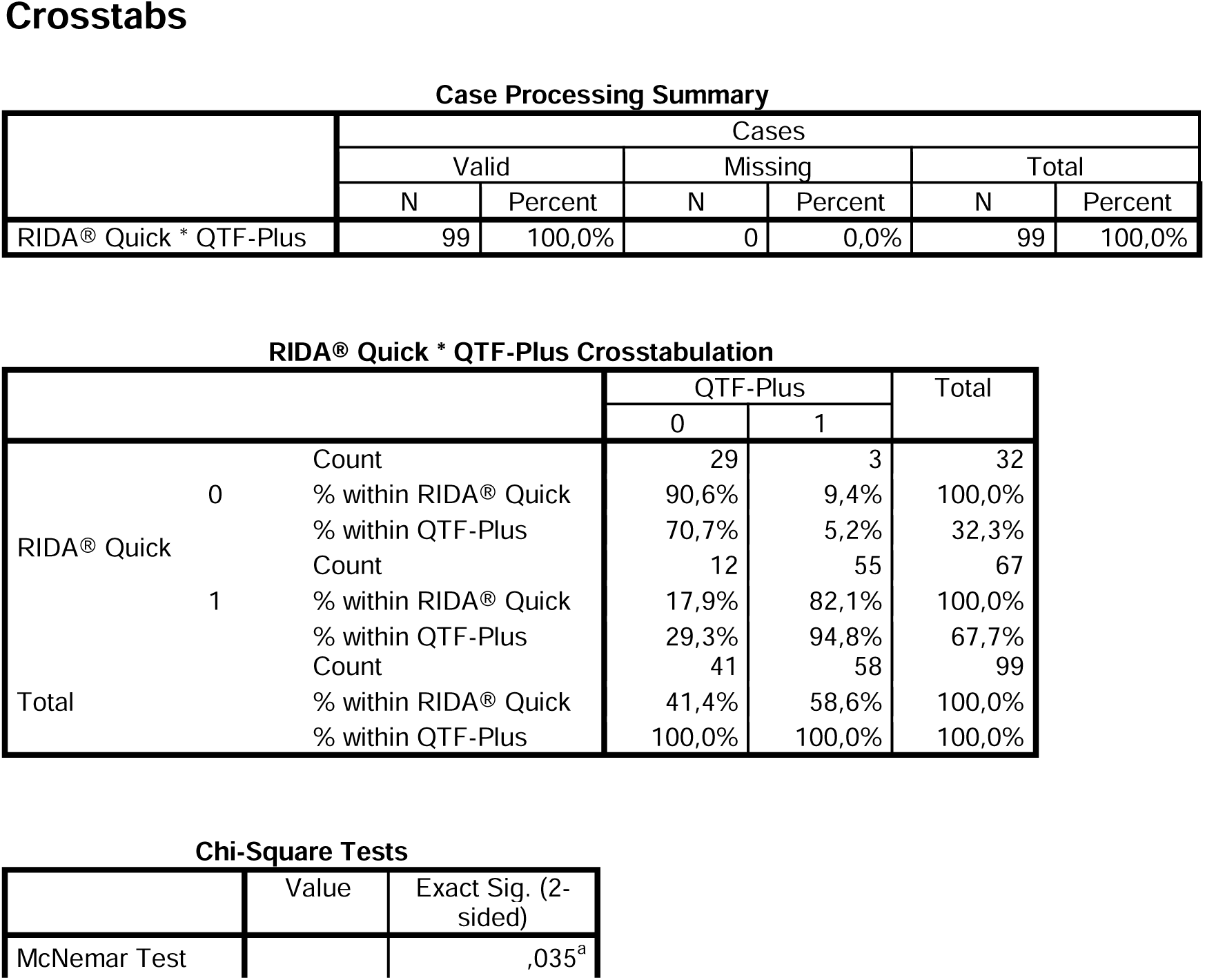

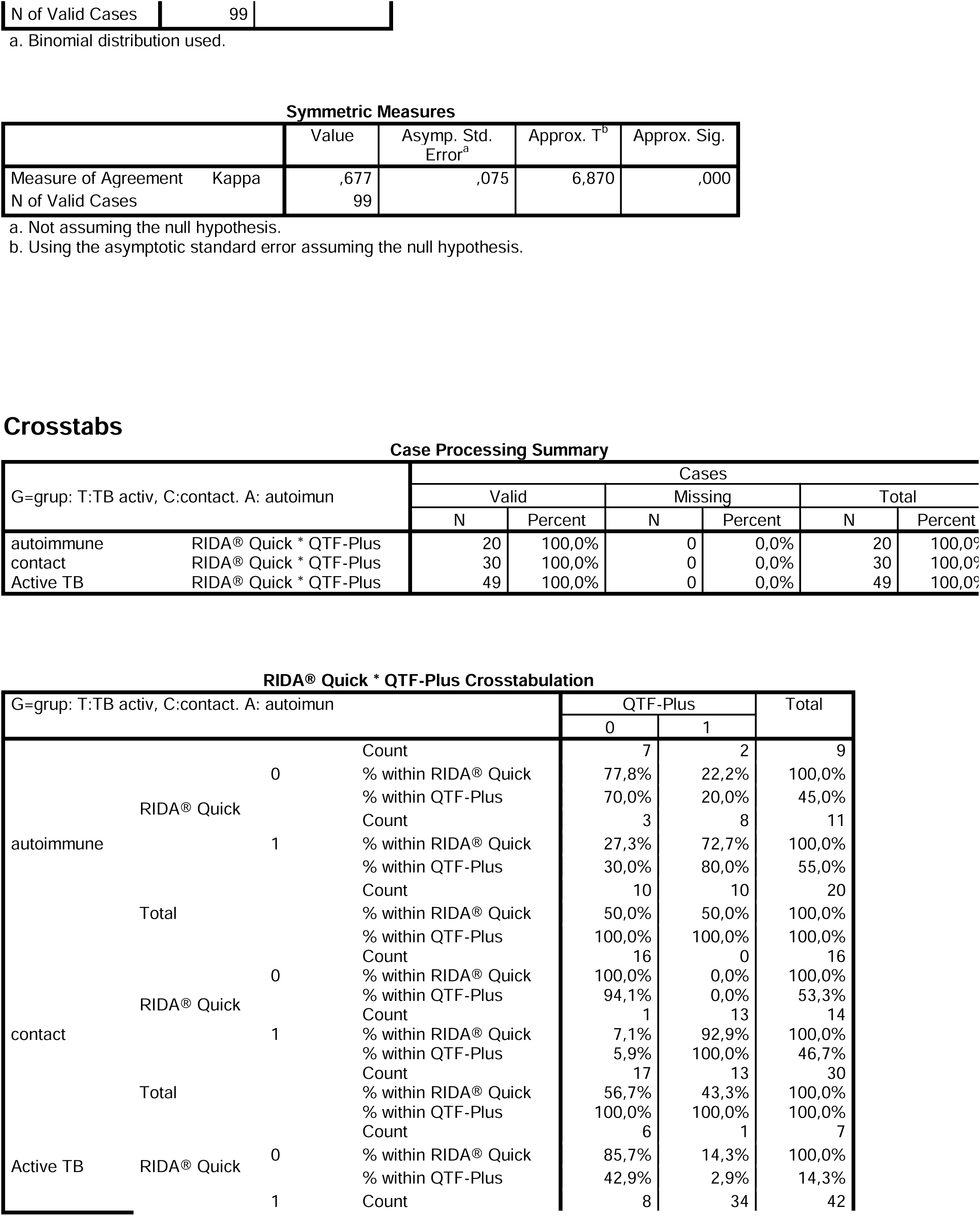

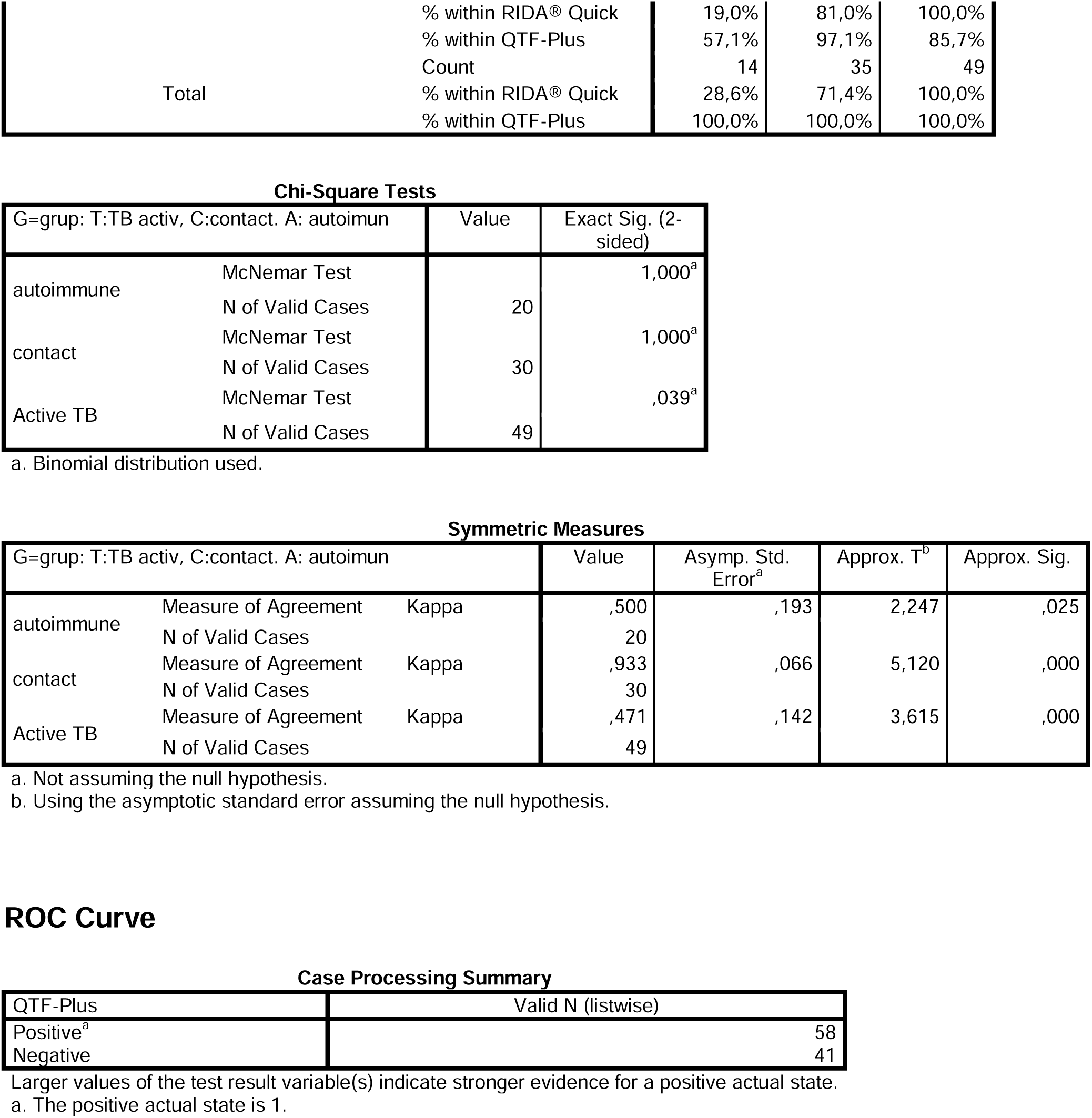

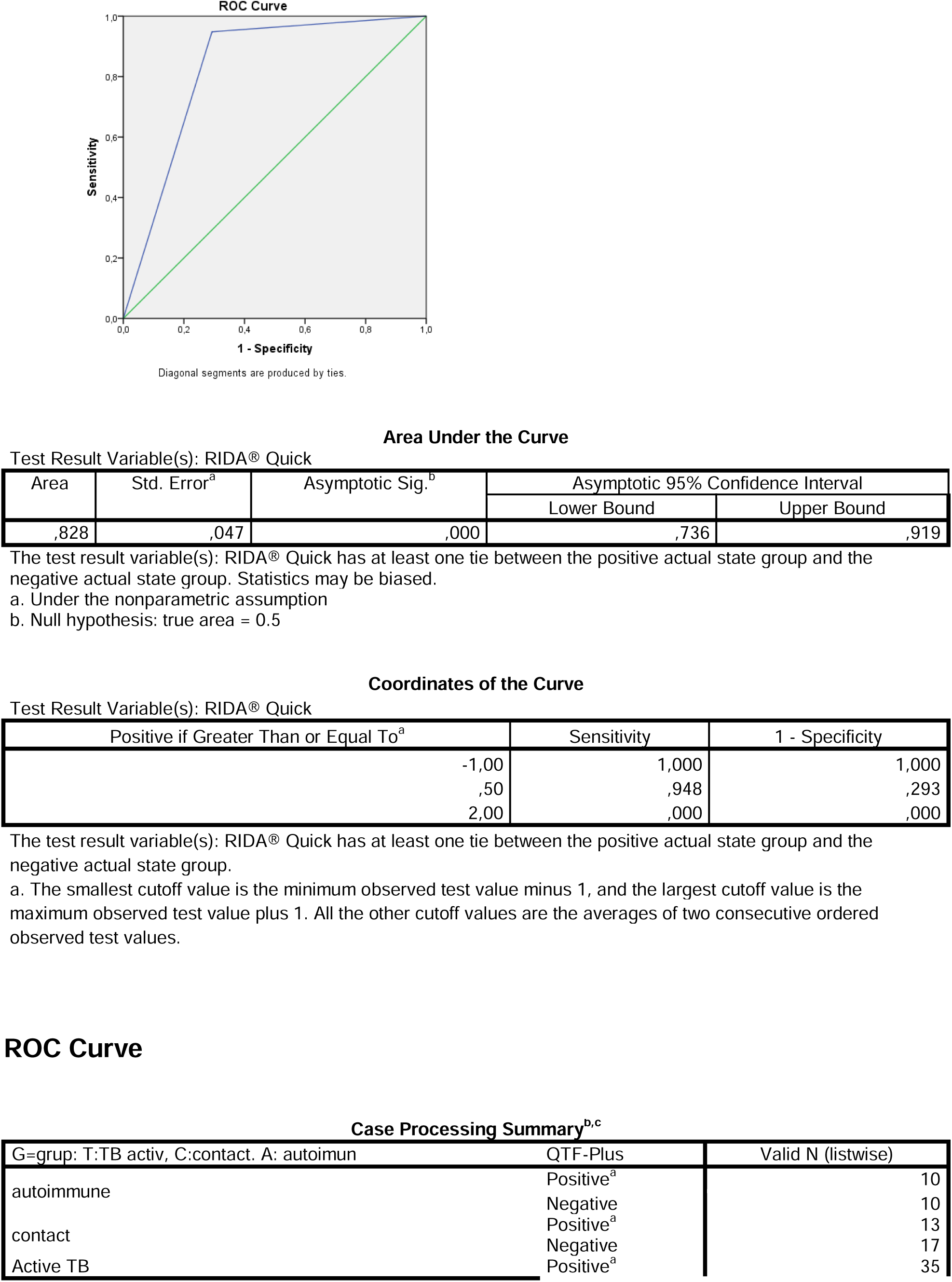

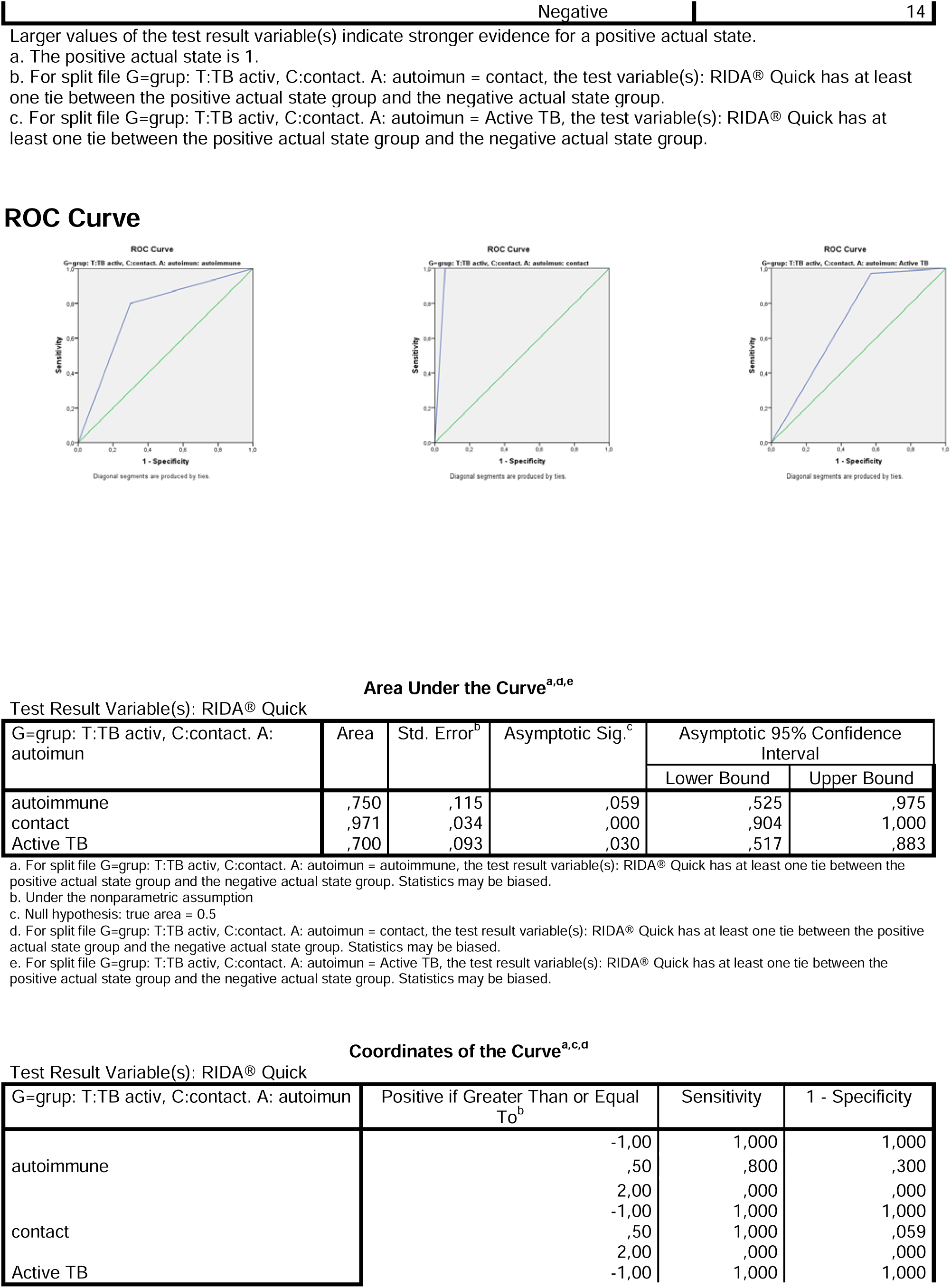

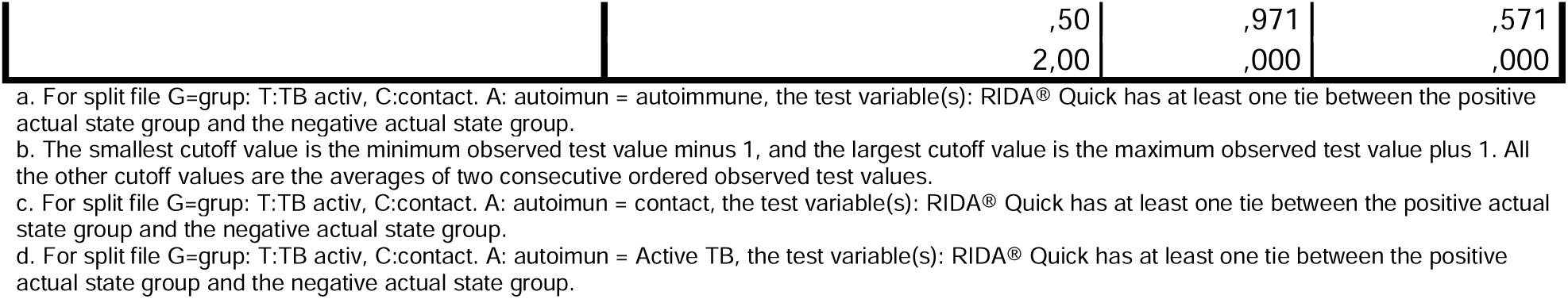

## ANNEX 2 RIDA®QUICK TB vs. TB laboratory assays

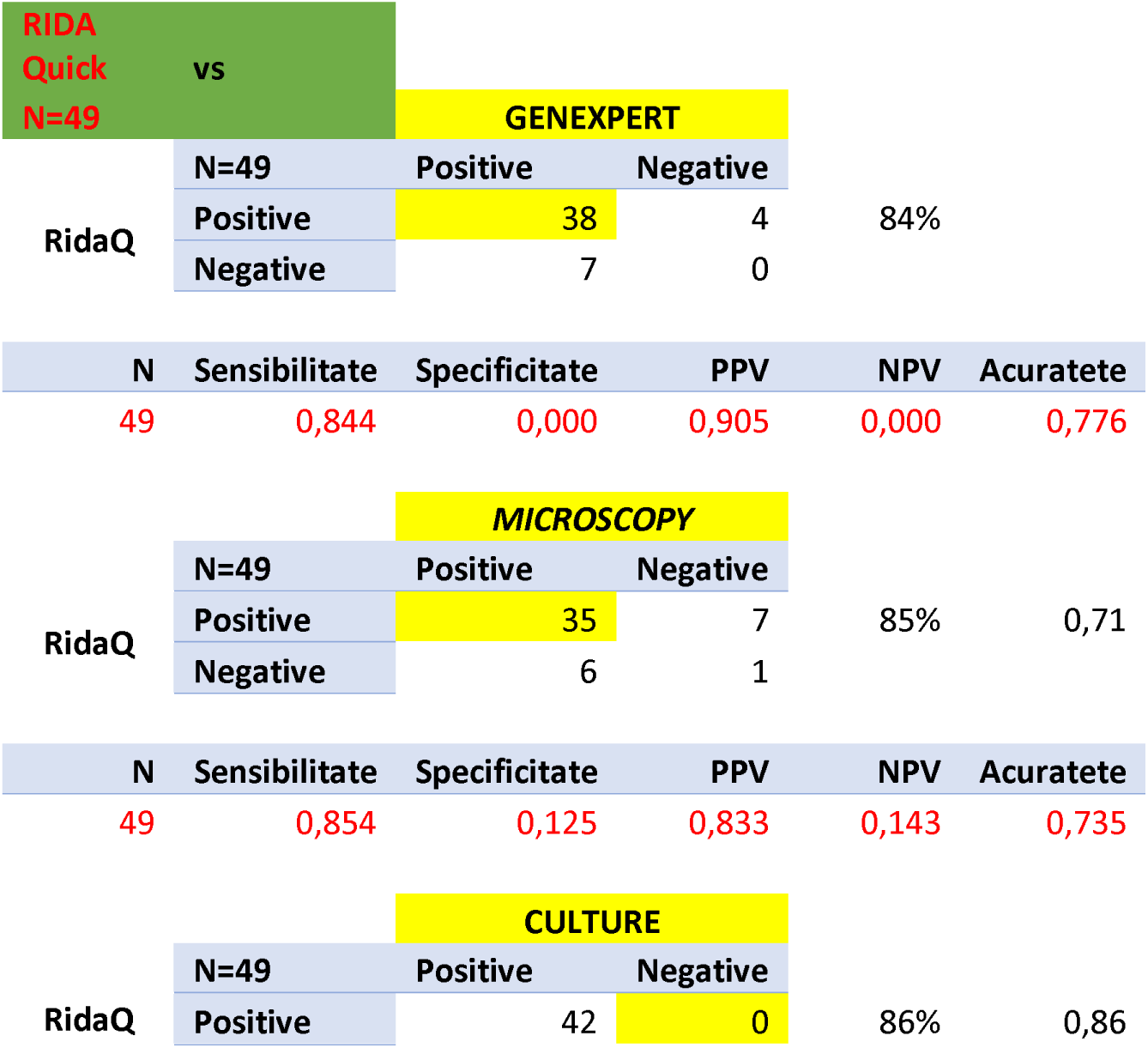
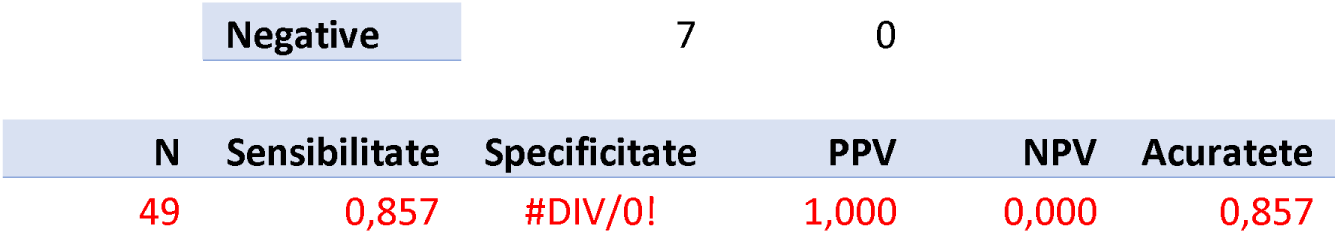

## ANNEX 3 RIDA®SCREEN TB vs QTF-Plus

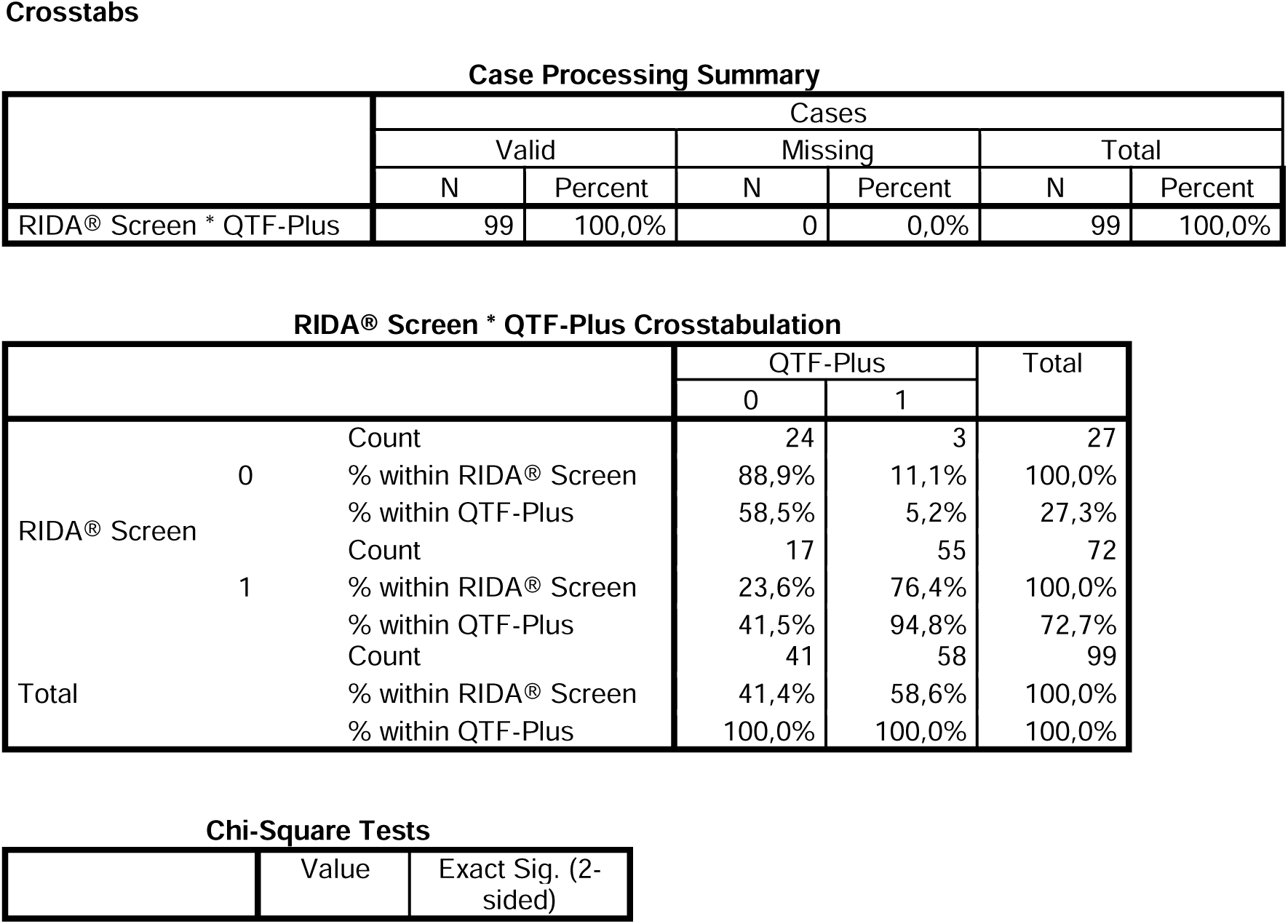

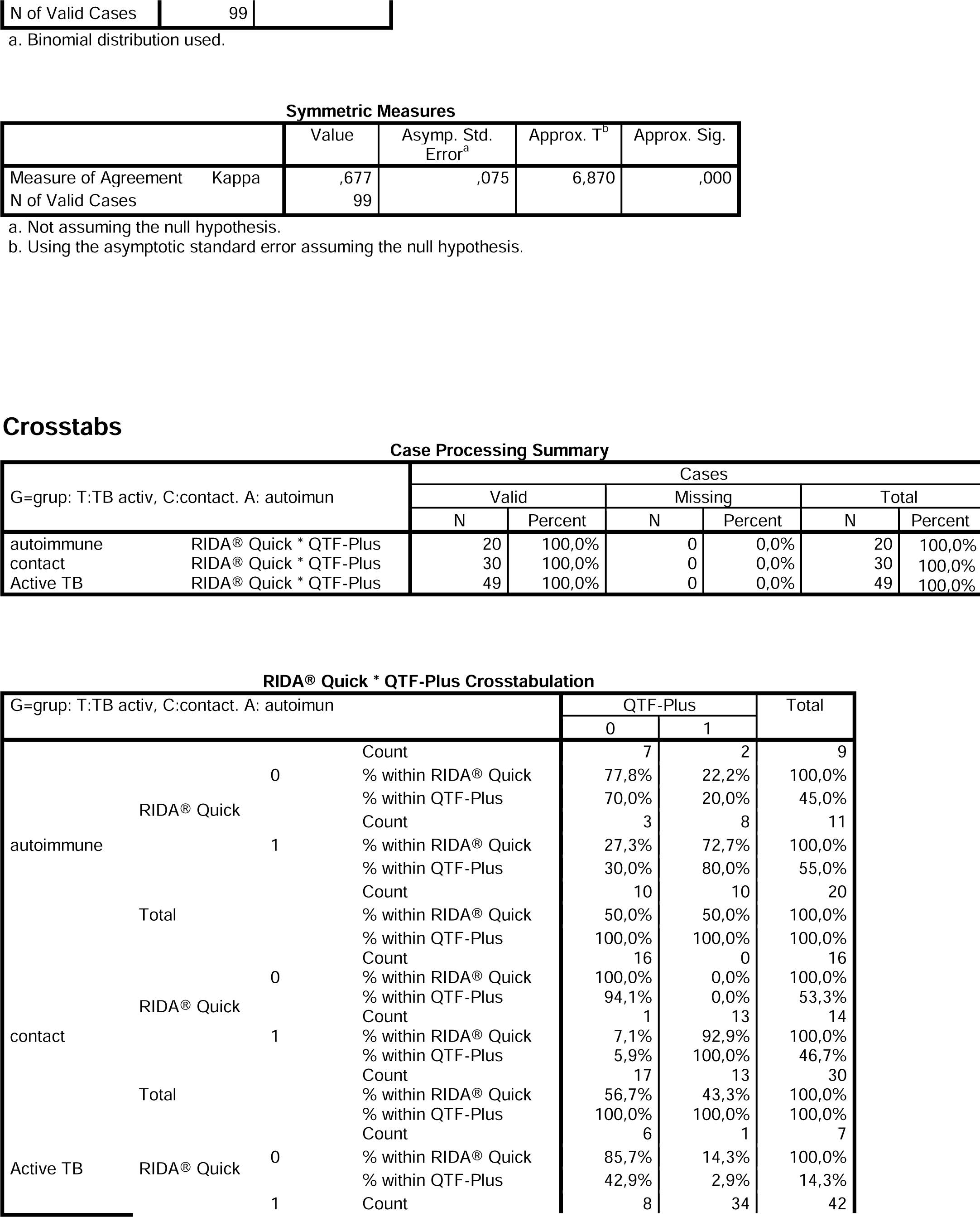

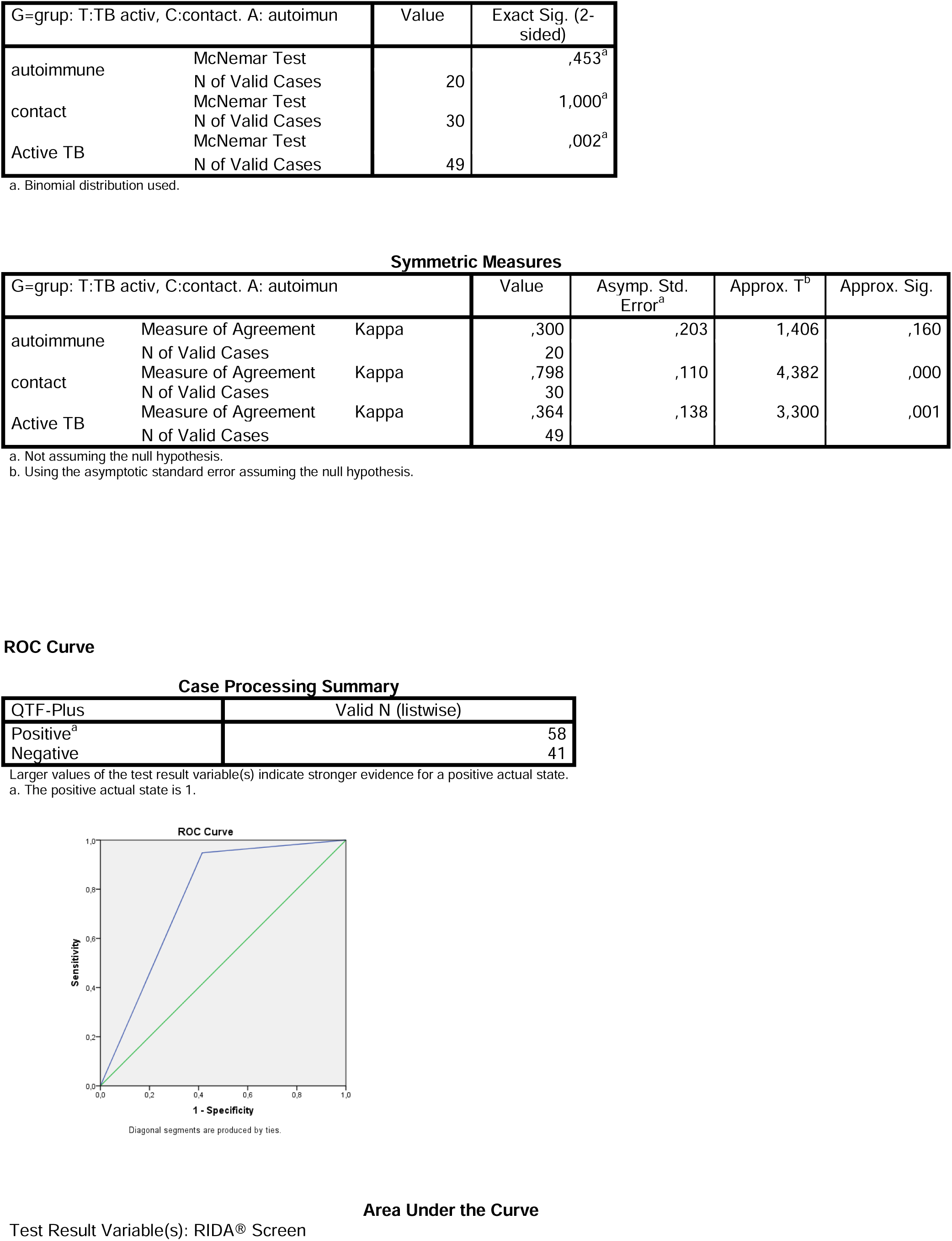

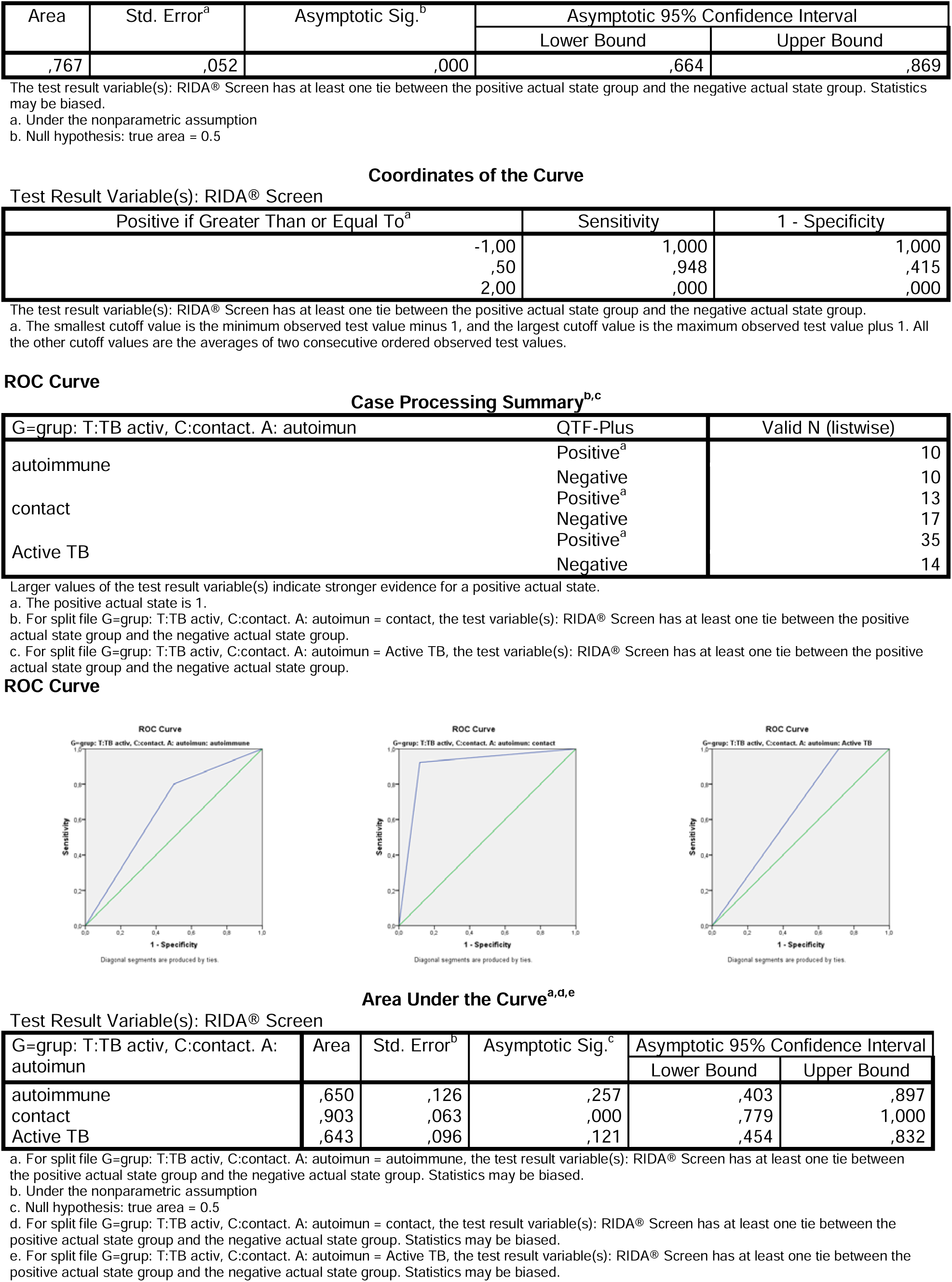

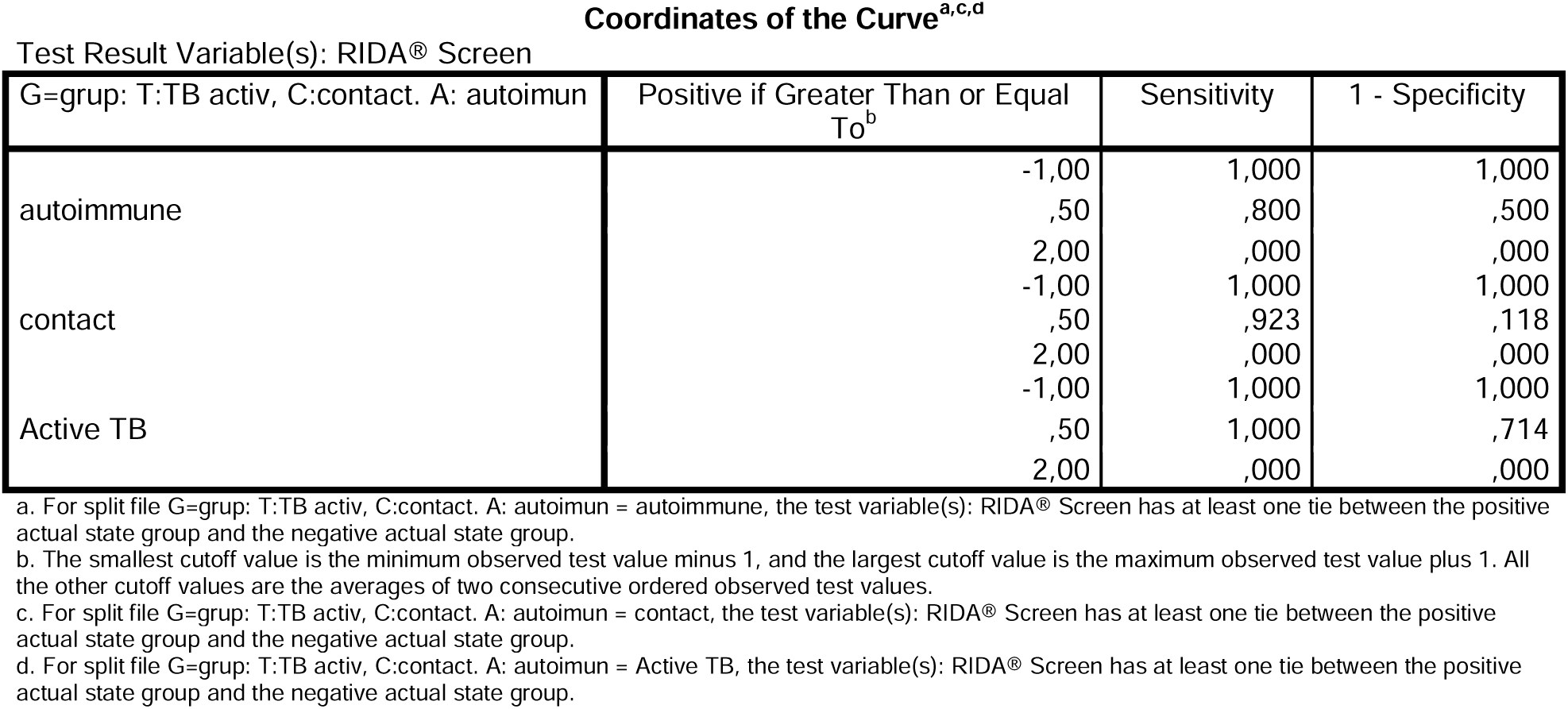

## ANNEX 4 RIDA®SCREEN TB vs. TB laboratory assays

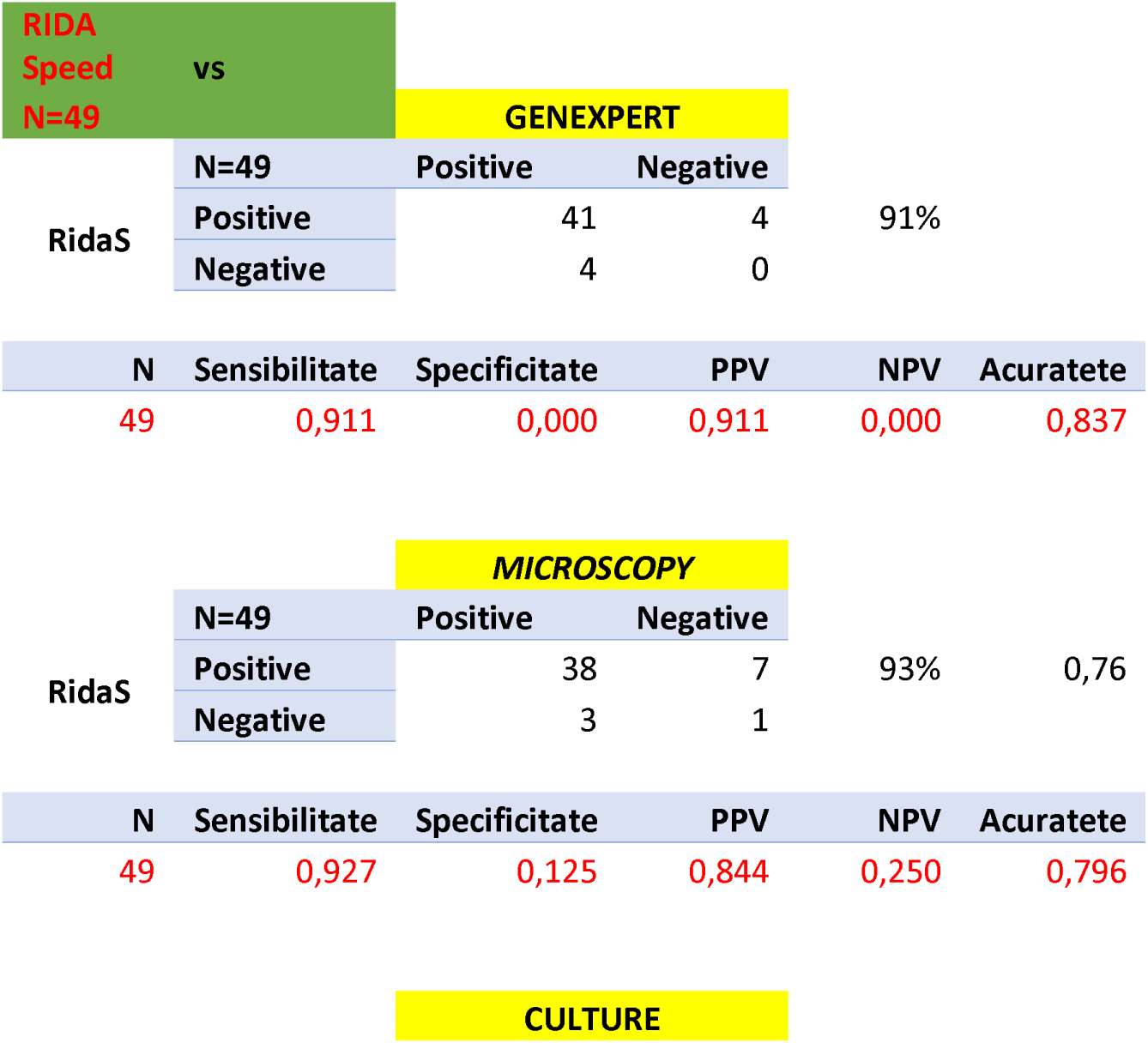
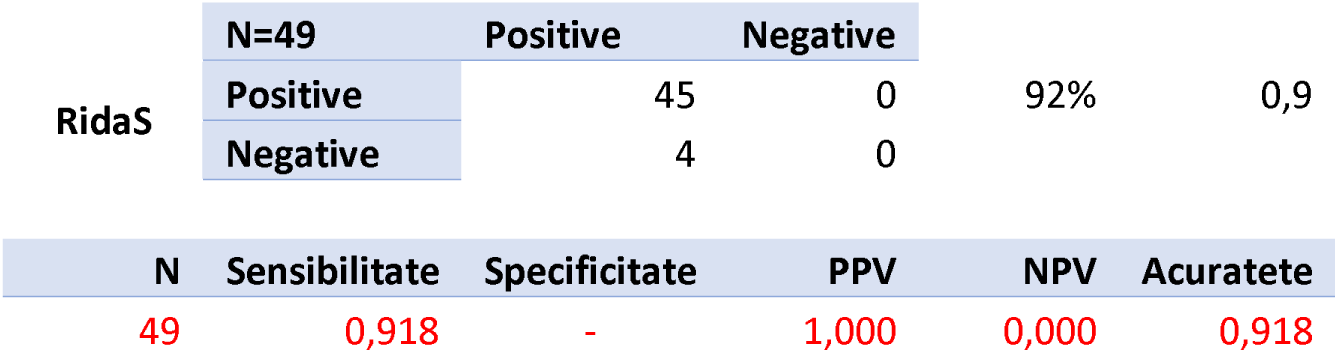

## ANNEX 5 QFT-Plus vs. TB laboratory assays

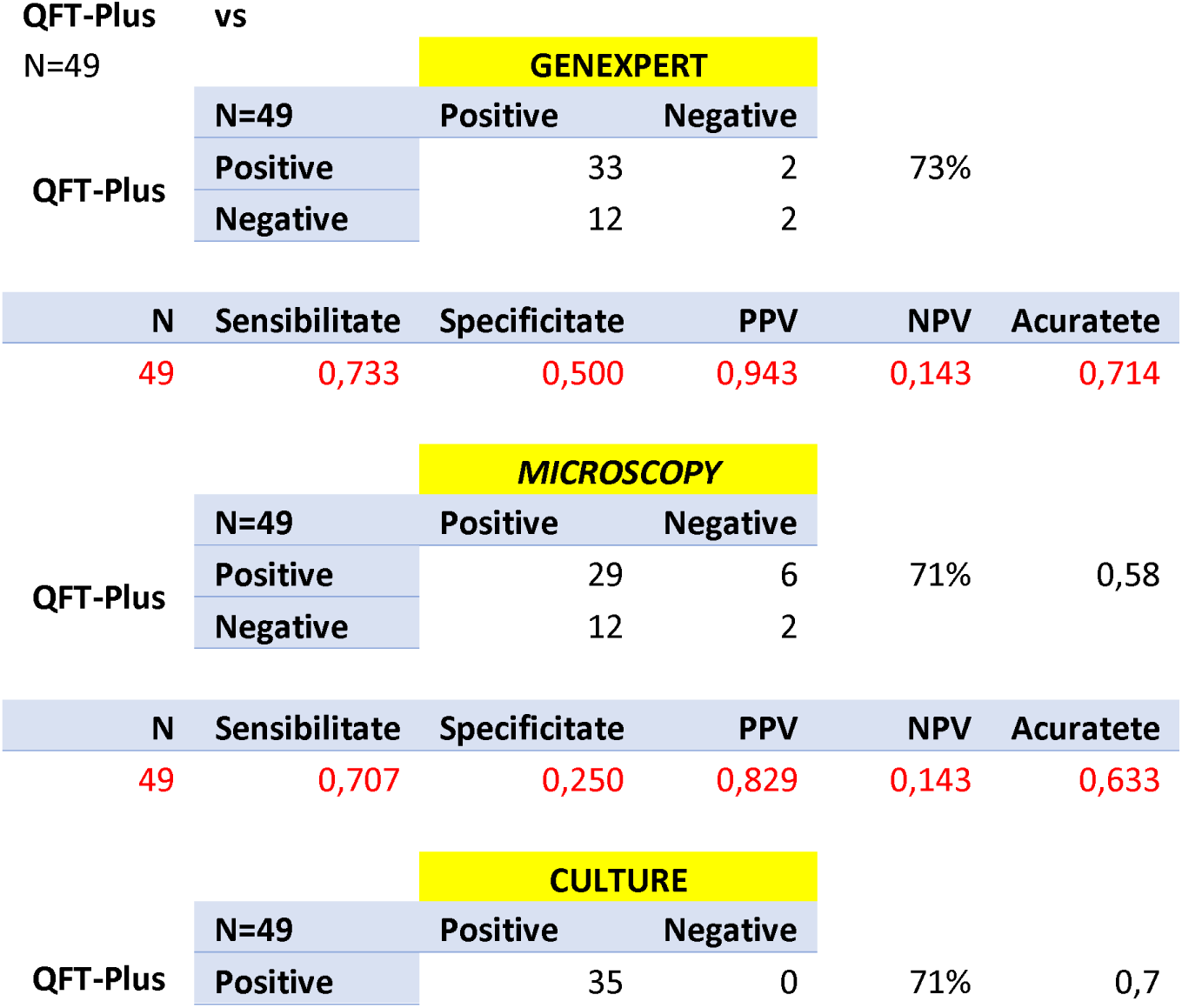
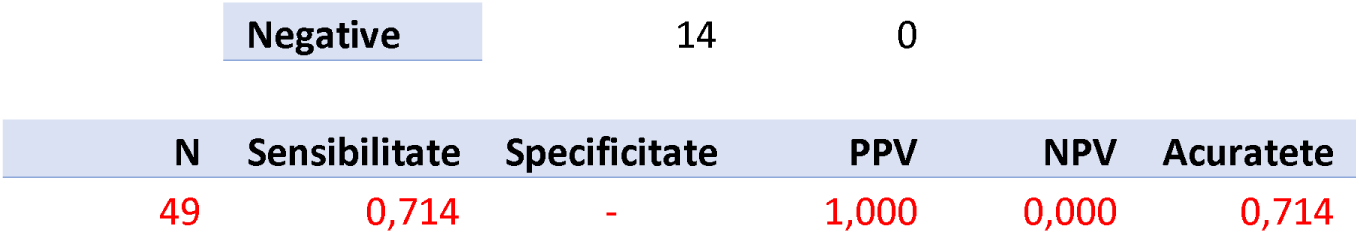

